# Nigro-striatal deficits capture phenoconversion risk in isolated REM sleep behavior disorder

**DOI:** 10.64898/2026.08.26.26361210

**Authors:** Martin Johansson, Alexis Baron, Rahul Gaurav, Anthony Ruze, Pauline Dodet, Aurélie Kas, Vineeth Radhakrishnan, Romain Valabrègue, Nicolas Villain, Graziella Mangone, Marie Vidailhet, Jean-Christophe Corvol, Isabelle Arnulf, Stéphane Lehéricy

**Author notes:** **Correspondence to**: Martin E. Johansson, **Full address**: Paris Brain Institute, Sorbonne Université, Pitié-Salpêtrière Hospital, 47 bd de, l’Hôpital, 75013, Paris, France, **Email:**.

## Abstract

Isolated rapid eye movement sleep behavior disorder (iRBD) is characterized by nigro-striatal deficits, comprising dopaminergic denervation of the striatum and loss of dopaminergic cells in the substantia nigra (SN), that may herald phenoconversion to clinically manifest synucleinopathy. While phenoconversion has repeatedly been shown to relate to pre-synaptic dopaminergic deficits in the striatum, potential involvement of loss of dopaminergic cells in the SN remain unclear. In addition, phenoconversion may independently relate to noradrenergic deficits, stemming from cell loss in the locus coeruleus/subcoeruleus (LC/LsC) complex.

Fifty-six iRBD patients were included and clinically followed over an 11-years as part of the ICEBERG study. Putamen dopamine denervation was quantified using ^123^I-FP-CIT single-photon emission computed tomography. Cell loss in the SN and LC/LsC was quantified using neuromelanin-sensitive magnetic resonance imaging (MRI). SN cell loss was additionally characterized as free water, derived from diffusion-weighted MRI. The primary outcome was time to phenoconversion. Cox proportional hazards regression was used to investigate relationships between phenoconversion risk and imaging predictors, estimated as hazard ratios (HRs).

Out of 56 patients, 24 (41%) converted to a clinically manifest synucleinopathy [PD=14 (58%), DLB=8 (33%), MSA=2 (8%)] over a maximum period of 11 years. We replicated the well-established finding that reduced putamen DaT confers an increased phenoconversion risk [HR (95%CI)=3.1 (1.7-5.5), P<0.001]. We extend on this by showing a similar relationship for SN neuromelanin [HR (95%CI)=2.5 [1.3-4.6], P=0.004], SN free water [HR (95%CI)=1.54 (1.06-2.24), P=0.025], and LC/LsC neuromelanin [HR (95%CI)=2.1 (1.2-3.7), P=0.011], demonstrating involvement of the broader nigro-striatal dopaminergic system along with potential involvement of noradrenergic neurotransmission. When adjusting for putamen DaT, the relationship between phenoconversion risk and SN neuromelanin was attenuated [P=0.16], suggesting partial overlap between the metrics. In contrast, when modelled together, SN neuromelanin [HR (95%CI)=2.8 (1.4-5.6), P=0.003] and LC/LsC neuromelanin [HR (95%CI)=2.3 (1.1-4.8), P=0.037] contributed to phenoconversion risk independently of each other, indicating a differential contribution of dopaminergic and noradrenergic neurotransmitter deficits to iRBD phenoconversion.

We demonstrate that phenoconversion in iRBD relates similarly to dopaminergic denervation of the putamen and cell loss in the SN. This opens possibilities for using NM-MRI, which can simultaneously capture dopaminergic and noradrenergic deficits, as an alternative to nuclear imaging techniques when estimating phenoconversion risk in iRBD.

## Introduction

Isolated REM sleep behaviour disorder (iRBD) is a parasomnia characterised by a loss of muscle atonia during REM sleep, resulting in dream-enactment and potentially violent movements.^1^ Within approximately 12 years of diagnosis, 75% of patients with iRBD phenoconvert to a clinically manifest synucleinopathy, including Parkinson’s disease, dementia with Lewy bodies, or multiple system atrophy.^2^ Such high phenoconversion rates position iRBD as a pre-symptomatic stage of synucleinopathy, offering insight into the underlying neural determinants of clinical progression and possibilities to devise interventions that modify their longitudinal trajectories.^3,4^ In this study, we investigated the relationship between phenoconversion risk in iRBD and nigro-striatal deficits, a core pathological hallmark of synucleinopathies.

iRBD is associated with deficits in the nigro-striatal system, comprising dopaminergic denervation of the striatum and cell loss in the substantia nigra (SN).^5^ Striatal dopamine denervation, as measured with nuclear imaging techniques,^6^ has emerged as a highly reliable indicator of phenoconversion risk in iRBD, pointing to a mechanistic role in the transition from pre-symptomatic to clinically overt disease.^2,7–11^ However, contributions of SN cell loss, which can be quantified using magnetic resonance imaging (MRI),^12–15^ remain less clear. Additionally, it is unknown whether SN cell loss and striatal dopamine denervation capture overlapping or independent variability in phenoconversion risk. This has implications for understanding the mechanisms whereby iRBD transitions into synucleinopathies, but it also matters practically: nuclear imaging of striatal dopamine is more costly and less widely available than MRI, and its involvement of radiation exposure complicates its use as an endpoint in clinical trials. MRI-based quantification of SN cell loss could provide an alternative marker of phenoconversion that overcomes these issues, permitting both low-burden stratification and subsequent longitudinal assessment of a well-defined imaging marker of disease progression.^16,17^

Although direct evidence from cohorts of iRBD patients is limited,^18^ research on the dynamics of nigro-striatal deficits in Parkinson’s disease supports a role for SN cell loss in phenoconversion. In Parkinson’s disease, both striatal dopamine denervation and SN cell loss precede the onset of motor symptoms by several years.^19–21^ However, at the time of diagnosis, the severity of striatal dopamine depletion typically exceeds the extent of SN cell loss.^22–24^ Consequently, while striatal dopamine denervation and SN cell loss progress largely in parallel, they also follow partly independent trajectories, which could result in differential contributions to phenoconversion risk in iRBD.

A potential relationship between SN cell loss and phenoconversion risk in iRBD may additionally be modulated by disturbances in other neurotransmitter systems, most notably by the noradrenergic system originating in the locus coeruleus (LC).^25,26^ In iRBD, dream-enacting behaviors reflect pathology in pontomedullary nuclei that gate motor inhibition during REM sleep.^27^ These nuclei include the subcoeruleus (LsC), a non-noradrenergic structure located immediately ventral to the LC, which is affected at an earlier stage of the pathological cascade that causes cell loss in the SN.^25,28,29^ Using neuromelanin (NM)-sensitive MRI, cell loss in the LC/LsC complex has been detected in iRBD^30–33^ and in clinically manifest synucleinopathies.^25,28,34,35^ Although the LC and LsC are largely indistinguishable in NM-MRI, evidence from nuclear imaging corroborates a related deficit in noradrenergic neurotransmission,^32^ which may sensitize dopaminergic SN cells to neurodegenerative processes and thereby hasten their decline.^26^ Taken together, this suggests that SN and LC/LsC integrity may carry both unique and complementary information about phenoconversion risk in iRBD.

In this study, we quantified nigro-striatal deficits using a combination of nuclear imaging and MRI in a single-center cohort of patients with iRBD (n=56) whose diagnoses were monitored over an 11-year period. Our principal aim was to establish how SN cell loss relates to phenoconversion risk in iRBD. In addition, we investigated the degree to which SN cell loss contributed to phenoconversion risk independently of striatal dopamine denervation and cell loss in the LC/LsC.

## Materials and methods

### Participants

58 patients with polysomnography-confirmed iRBD^36,37^ from the longitudinal ICEBERG study (clinicaltrials.gov identifier: NCT02305147) were included. Inclusion criteria were age ≥18 to ≤75 years, ≤4 years disease duration, with minimal or no cognitive disturbances (Mini-Mental State Examination score>26). At baseline, patients underwent clinical assessments, MRI scanning, and ^123^I-FP-CIT (DATSCAN®) single-photon emission computed tomography (SPECT) at the Paris Brain Institute (Sorbonne Université, Pitié-Salpêtrière Hospital). Diagnoses were monitored longitudinally during yearly visits to the sleep clinic between November 2015 and February 2026. Conversions to Parkinson’s disease,^38^ dementia with Lewy bodies,^39^ or multiple system atrophy^40^ were established in accordance with international criteria. Patients who converted to non-synucleinopathies were excluded (n=1, Alzheimer’s disease), along with patients lacking longitudinal follow-up data (n=1). An independent sample of 165 patients with Parkinson’s disease and 61 healthy controls from ICEBERG were additionally included to derive reference values for standardization of imaging metrics and to compute disease-related cut-offs for imaging metrics, enabling stratification of iRBD patients into normal and abnormal groups. Research was sponsored by Inserm, conducted in accordance with Good Clinical Practice, and approved by the French regulation authorities and an ethical committee (IRB: 2014-A00725-42/48-12). Written informed consent was obtained for all participants.

### Image acquisition

#### Single-photon emission computed tomography

^123^I-FP-CIT SPECT acquisition is detailed elsewhere.^6^ Briefly, they were acquired using a Discovery NM/CT 670 Pro system 2-head imager (GE Healthcare). Acquisition was performed 3-4 hours after tracer injection. Scans were reconstructed on a GE Healthcare Xeleris Workstation using an iterative reconstruction algorithm that includes motion detection and correction, followed by post-filtering using a fourth-order low-pass filter with a cut-off frequency of 0.35cm^-1^. Lastly, correction for signal attenuation due to skull density was performed using the Chang method (μ=0.12 cm^-1^).

#### Magnetic resonance imaging

MRI scanning was performed using a Siemens 3T MAGNETOM Prisma Fit system equipped with a 64-channel head coil. Whole-brain structural scans were acquired using T1-weighted three-dimensional (3D) magnetization-prepared two rapid gradient echo (MP2RAGE) [TR/TE=5000ms/2.98ms, TI=700ms/2500ms, flip angle=4° and 5°, phase-encoding direction=A>P, voxel size=1.0×1.0×1.0mm^3^, field-of-view dimensions=176×232×256mm^3^]. NM-sensitive scans were acquired using a T1-weighted two-dimensional (2D) turbo spin echo (TSE) sequence, with a field-of-view oriented perpendicular to the longitudinal axis of the brainstem [TR/TE=890ms/13ms, flip angle=90° and 180°, phase-encoding direction=A>P, voxel size=0.43×0.43×3mm^3^, field-of-view dimensions=200×220×48mm^3^]. Diffusion-weighted scans were acquired using a multishell echo-planar imaging sequence [TR/TE=10400/59ms, flip angle=90°, scheme=monopolar, b-values=0-300-700-2000/mm^2^, phase-encoding direction=A>P, voxel-size=1.7×1.7×1.7mm^3^, field-of-view dimensions=128×128×84mm^3^, diffusion gradient directions=114, with 14 b0 images]. For diffusion-weighted scans, field maps consisting of single b0 images with an inverted phase-encoding direction (P>A) were acquired to correct for susceptibility distortions during image preprocessing.

### Image preprocessing

#### Single-photon emission computed tomography

Putamen dopamine was quantified using ^123^I-FP-CIT SPECT scans. The quantification is detailed elsewhere.^6^ Briefly, SPECT scans were rigidly aligned to their corresponding T1-weighted MRIs. T1-weighted images underwent tissue-type and subcortical segmentation using the FMRIB Software Library (FSL)^41^ FAST^42^ and FIRST^43^ workflows, respectively. Tissue-type segmentations of cerebrospinal fluid, white matter, and three subdivisions of grey matter based on dopamine density (high=[putamen, caudate, accumbens]; intermediate=pallidum; low=[cortex, thalamus]) were used to correct for partial volume effects using Yang’s iterative region-based voxel-wise method (30 iterations; convergence visually verified for all participants).^6,44^ DaT-specific binding ratios (SBR) were calculated for the bilateral putamen signal relative to the bilateral occipital cortex.

### Magnetic resonance imaging

#### Substantia nigra and locus (sub)coeruleus neuromelanin

Degeneration of the SN and LC/LsC was quantified using NM-sensitive MRI. Fully automated segmentations of the SN and LC were obtained in accordance with previously described work.^13,30,31,33,45,46^ For the SN, automated segmentation was performed using NigraNet, a deep learning approach based on convolutional neural networks, which defines regions of interest in the SN and a background reference region containing the tegmentum and superior cerebellar peduncles.^45^ For the LC/LsC, regions of interest were placed in the pons.^31,33^ The LC/LsC was then defined within this region by extracting 10 connected voxels with maximum signal intensity. A background reference region was defined in the rostral pontomesencephalic area. For both SN and LC/LsC, signal-to-noise ratios (SNR) were calculated bilaterally as the mean NM signal in the region of interest divided by the NM signal obtained from the corresponding background reference region, multiplied by 100.^47^

#### Substantia nigra free water

SN degeneration was additionally quantified using diffusion-weighted MRI. Quantification of free water (FW) content in the SN followed well-established pipelines.^48–51^ Diffusion-weighted images were preprocessed using QSIPrep (version 1.0.1).^52^ This involved denoising through Marchenko-Pastur principal component analysis^53^ and Gibbs unringing,^54^ followed by corrections for head motion, eddy currents and susceptibility distortion.^55–58^ Lastly, images were resampled to 2 mm isotropic voxels in ACPC space. DiPY^59^ was used to compute images of mean b0, fractional anisotropy (FA), and FW. A study-specific FA template was created using antsMultivariateTemplateConstruction,^60–62^ with the HCP165_FA_2mm template in MNI152NLin6Asym-space included in FSL as the target.^48^ Non-linear transformations between the participant-specific FA images and the study-specific FA template were estimated using ANTs.^61^ These transformations were subsequently used to normalize b0 and FW images. In template-space, b0 images from all participants were averaged and used to manually draw a 4×4×4mm^3^ region of interest in the posterior SN of each hemisphere.^48,49^ Each region of interest was drawn along two horizontal slices, starting from the slice located directly below the most inferior portion of the red nucleus, where the SN appears hypointense. The posterior SN region of interest was subsequently used to extract bilateral SN FW values for each participant. In all analyses and in the calculation of composite scores (see below), SN FW sign was reversed to match the other imaging predictors.

### Calculation of composite scores

Composite scores were calculated to explore potential advantages of combining imaging predictors. First, raw imaging metrics were z-scored relative to reference values derived from patients with Parkinson’s disease according to the following formula

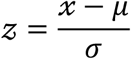

where *x* is the raw imaging metric of an individual iRBD patient, *μ* is the mean of patients with Parkinson’s disease, and *σ* is the standard deviation of patients with Parkinson’s disease. Z-scoring was performed separately for each imaging metric, using all available values. To characterize broader deficits in the nigro-striatal system, putamen DaT SBR was separately combined with SN NM SNR (Putamen DaT+SN NM) and SN FW (Putamen DaT+SN FW). To characterize the joint contribution of dopaminergic and noradrenergic degeneration, SN and LC/LsC NM SNR were combined (NM [SN+LC/LsC]).

### Statistical analysis

All analyses were performed in R (R Core Team 2025; version 4.5.2). Summary statistics report number (percentage) for categorical data and mean (standard deviation) for continuous data. Years from first assessment to phenoconversion or censoring were considered as the primary outcome. Kaplan-Meier survival analysis was used to estimate overall time to phenoconversion. Cox proportional hazards regression was used to test associations between time to phenoconversion and continuous imaging predictors, estimated as hazard ratios (HRs).^63,64^ To facilitate comparisons of HRs between predictors, all imaging metrics (including composite scores) were z-scored directly prior to each Cox regression analysis. Consequently, HRs represent changes in phenoconversion hazard associated with one standard deviation decrease per imaging predictor. Apparent discriminative performance was explored using concordance survival scores (Harrell’s C-index),^65^ time-dependent receiver-operating characteristic (ROC) area under the curve (AUC),^66,67^ and integrated ROC AUC (iAUC) calculated to maximum follow-up (11 years).^68^ Censoring was assumed to be uninformative. The proportional hazards assumption was verified statistically and through visual inspection of scaled Schoenfeld residuals plotted against time. Age and sex were considered as covariates of non-interest but showed no relationship with phenoconversion hazard (both P>0.5) and were therefore dropped to preserve statistical power. Sensitivity analyses to account for between-metric differences in data missingness were conducted on complete cases (i.e. only participants with all four imaging metrics) and following multiple imputation^69,70^ (see Supplementary Materials).

Post hoc analyses were performed to address predictor-and disease-specificity. Cox regression analyses were performed specifically to assess if SN NM explained unique variability in phenoconversion hazard relative to other imaging predictors. In these analyses, SN NM was included as the main predictor and its interaction with either putamen DaT, SN FW was assessed. Additionally, diagnosis-specific Cox regression was used to test for associations between imaging predictors and Parkinson’s disease-specific phenoconversion risk. In these analyses, conversions to diagnoses of non-interest (i.e., dementia with Lewy bodies and multiple system atrophy) were treated as censoring events and hence considered uninformative. Due to limited sample sizes, this analysis was not performed for dementia with Lewy bodies and multiple system atrophy.

A secondary Cox regression analysis was performed to assess the utility of stratifying iRBD patients using imaging-based abnormality classifications. For each imaging predictor, disease-related abnormality cut-offs were derived from an independent cohort of Parkinson’s disease patients (N=165) and healthy controls (N=61) who participated in the ICEBERG study. Binomial logistic regression followed by ROC curve analysis was used to determine how well imaging predictors could distinguish Parkinson’s disease patients from controls. Youden’s J-statistic was used to determine the cut-off point with optimal discrimination performance. Cut-offs were subsequently used to classify iRBD patients as either abnormal or normal. Cox regression was then used to assess whether imaging-based abnormalities captured phenoconversion risk.

## Results

### Characterization of overall phenoconversion risk

Demographic and clinical information can be found in **Table 1**. Out of 56 iRBD patients, 24 (43%) converted to a clinically manifest synucleinopathy [Parkinson’s disease=14 (58%), dementia with Lewy bodies=8 (33%), multiple system atrophy=2 (8%)] over a maximum follow-up period of 11 years (**Table 1**; **Figure 1A-B**). The median disease-free time was 9.0 years, with a restricted mean disease-free time of 7.7 years (upper limit: 10.3 years). Kaplan-Meier survival analysis showed that phenoconversion risk was 15% at year 2 [95% CI=5%-23%], 18% at year 4 [95% CI=7%-28%], 25% at year 6 [95% CI=12%-35%], 35% at year 8 [95% CI=20%-48%], and 64% at year 10 [95% CI=38%-79%]. The overall phenoconversion rate per year was 6.9%. Notably, the estimated hazard function indicated that phenoconversion rates remained relatively constant until the four-year mark, after which they accelerated linearly (**Figure 1B**).

**Table 1.** Demographics and baseline characteristics. PD=Parkinson’s disease; DLB=Dementia with Lewy bodies; MSA=Multiple system atrophy; M=Male; F=Female; MDS-UPDRS=Movement Disorders Society-sponsored Unified Parkinson’s Disease Rating Scale; MoCA=Montreal Cognitive Assessment; UPSIT= University of Pennsylvania Smell Identification Test; SCOPA-AUT=Scales for Outcomes in Parkinson’s Disease – Autonomic Dysfunction; SN=Substantia nigra; NM=Neuromelanin; FW=Free water; LC/LsC=Locus coeruleus/subcoeruleus; NA=Not available.

| Variable | N | Overall<br>N = 56<sup>1</sup> | Phenoconversion status |  |  |  |
| --- | --- | --- | --- | --- | --- | --- |
|  |  |  | Non-converter<br>N = 32<sup>1</sup> | PD<br>N = 14<sup>1</sup> | DLB<br>N = 8<sup>1</sup> | MSA<br>N = 2<sup>1</sup> |
| Age at inclusion | 56 | 67.8 (5.7) | 67.9 (4.8) | 68.3 (7.2) | 68.3 (5.9) | 61.0 (5.7) |
| Sex | 56 |  |  |  |  |  |
| M |  | 50 (89%) | 29 (91%) | 12 (86%) | 7 (88%) | 2 (100%) |
| F |  | 6 (11%) | 3 (9%) | 2 (14%) | 1 (13%) | 0 (0%) |
| Duration since iRBD diagnosis (years) | 56 | 1.8 (3.2) | 2.3 (3.9) | 1.3 (1.9) | 1.1 (1.8) | 1.6 (0.1) |
| Time to last follow-up (years) | 56 | 6.2 (2.9) | 6.9 (2.5) | 4.8 (3.4) | 6.8 (2.4) | 1.5 (0.3) |
| MDS-UPDRS part III (OFF) | 56 | 11.0 (6.0) | 10.3 (6.1) | 11.6 (5.6) | 13.8 (6.3) | 7.5 (4.9) |
| MoCA | 56 | 27.3 (2.3) | 27.8 (2.0) | 27.4 (2.1) | 25.5 (3.3) | 28.0 (1.4) |
| UPSIT | 54 | 20.2 (6.0) | 20.1 (6.1) | 19.1 (5.2) | 21.3 (3.9) | 24.5 (16.3) |
| SCOPA-AUT | 56 | 13.1 (6.2) | 12.6 (6.6) | 14.4 (6.1) | 12.9 (6.1) | 13.5 (2.1) |
| Putamen DaT | 41 | 3.3 (0.6) | 3.6 (0.6) | 3.1 (0.6) | 3.1 (0.6) | 2.2 (0.4) |
| SN NM | 40 | 112.0 (2.1) | 112.8 (2.1) | 110.9 (1.3) | 111.8 (1.9) | 108.6 (NA) |
| SN FW | 46 | 0.25 (0.06) | 0.23 (0.04) | 0.28 (0.06) | 0.28 (0.10) | 0.20 (NA) |
| LC/LsC NM | 46 | 124.1 (3.8) | 124.9 (3.7) | 123.2 (3.8) | 123.1 (4.3) | 122.9 (NA) |

<sup>1</sup>Mean (SD) for continuous variables or Frequency (%) for nominal variables.

**Figure 1.**
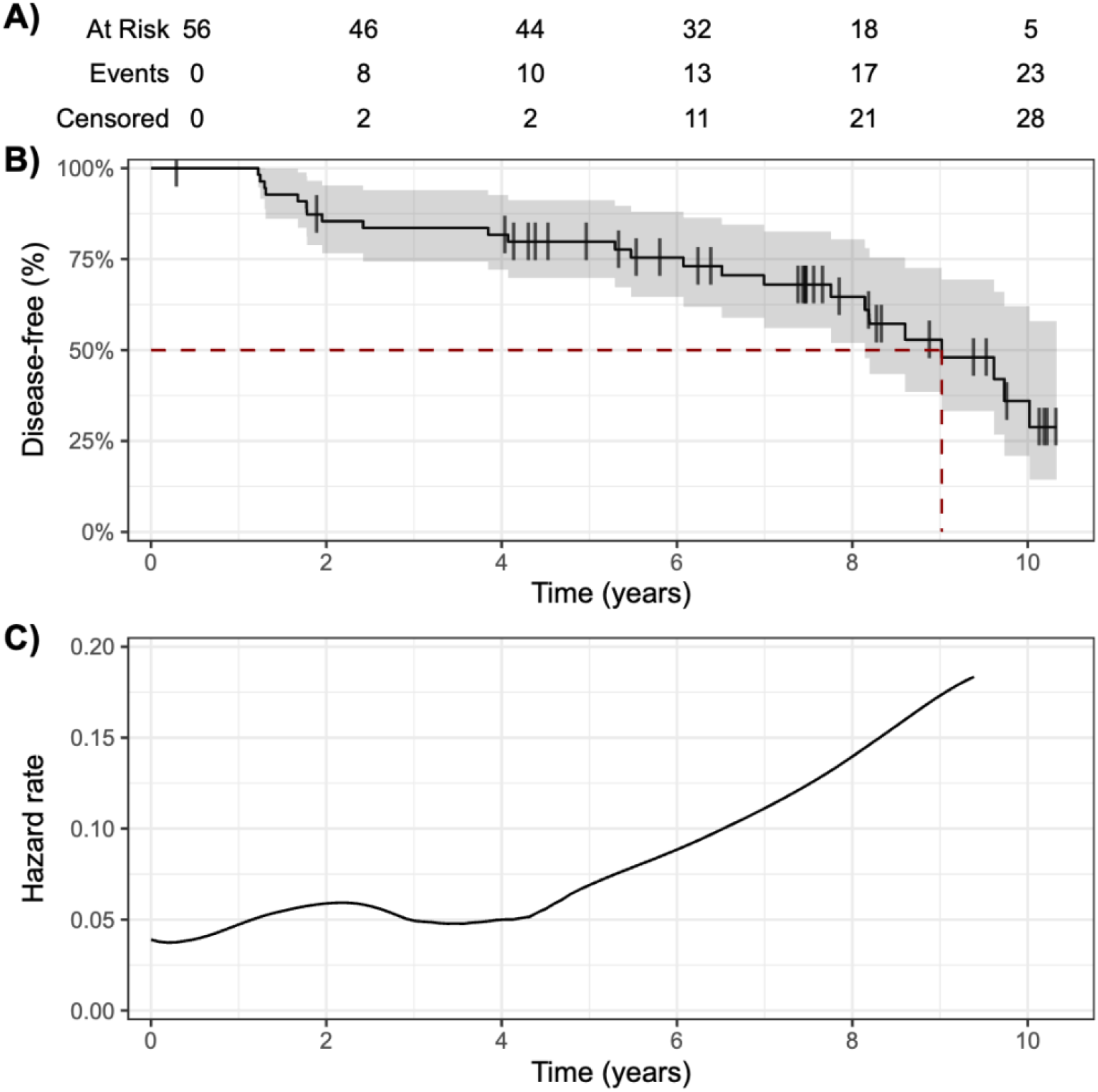
Phenoconversion in iRBD. (A) Risk-table showing numbers of phenoconversions and censoring events over time. (B) Estimated probability of remaining disease-free, with median disease-free time highlighted (red dashed line). (C) Estimated hazard rate as a function of time.

### Putamen DaT and substantia nigra neuromelanin capture similar variability in phenoconversion risk

Lower putamen DaT was associated with higher phenoconversion risk [**Table 2**; **Figure 2A**; HR (95%CI)=3.1 (1.7-5.5), χ^2^(1)=14.7, P<0.001]. Putamen DaT showed excellent performance in ranking patients based on time to phenoconversion [**Figure 2B**; C-index=0.81] and in identifying converters [**Figure 2C-D**; iAUC=0.83]. Lower SN NM was similarly associated with higher phenoconversion risk [HR (95%CI)=2.5 [1.3-4.6], χ^2^(1)=8.1, P=0.004]. SN NM showed good performance in ranking patients based on time to phenoconversion [C-index=0.74] and in discriminating converters from non-converters [iAUC=0.76].

**Table 2.** Results from Cox proportional hazards regression with continuous imaging predictors. HR=Hazard ratio; CI=Confidence interval; iAUC=Integrated area under curve; SN=Substantia nigra; NM=Neuromelanin; FW=Free water; LC/LsC=Locus coeruleus/subcoeruleus.

| Predictor | N | % converters | HR (95% CI)<sup>1</sup> | $\chi^2(1)$ | P-value | C-index | iAUC |
| --- | --- | --- | --- | --- | --- | --- | --- |
| Putamen DaT | 41 | 0.46 | 3.1 (1.7-5.5) | 14.7 | <0.001 | 0.81 | 0.83 |
| SN NM | 40 | 0.45 | 2.5 (1.3-4.6) | 8.1 | 0.004 | 0.74 | 0.76 |
| SN FW | 46 | 0.43 | 1.54 (1.06-2.24) | 5.1 | 0.025 | 0.66 | 0.69 |
| LC/LsC NM | 46 | 0.43 | 2.1 (1.2-3.7) | 6.5 | 0.011 | 0.63 | 0.68 |
| SN NM+DaT | 29 | 0.52 | 3.4 (1.7-6.7) | 12.5 | <0.001 | 0.81 | 0.85 |
| SN FW+DaT | 33 | 0.48 | 2.3 (1.4-3.7) | 10.5 | 0.001 | 0.77 | 0.80 |
| NM (SN+LC/LsC) | 40 | 0.45 | 3.0 (1.7-5.5) | 13.5 | <0.001 | 0.77 | 0.81 |

<sup>1</sup>HRs reflect phenoconversion risk associated with one standard deviation decrease in a predictor.

**Figure 2.**
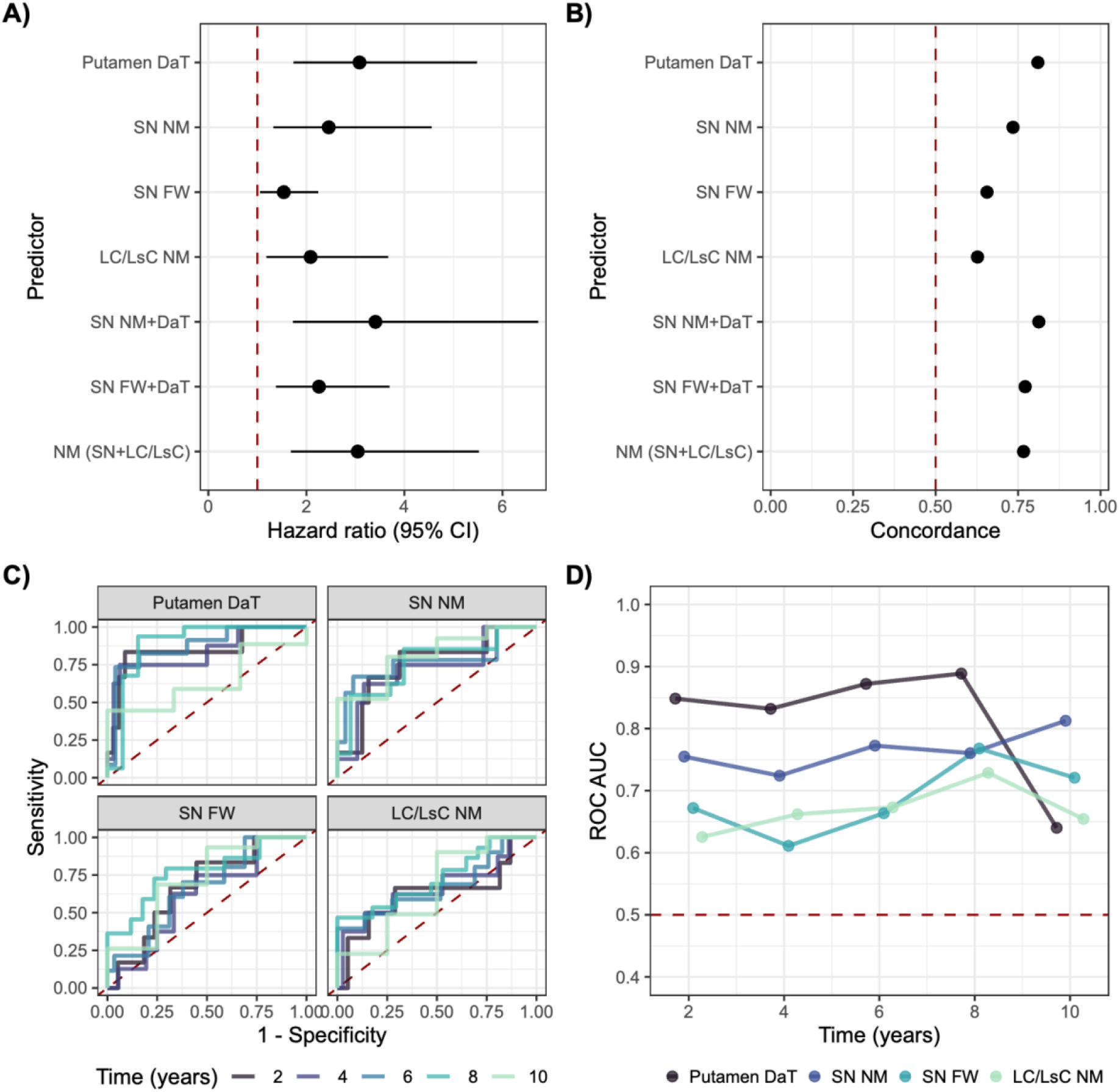
(A) Hazard ratios and 95% confidence intervals for imaging predictors. (B) apparent concordance (C-index) scores. (C) Apparent time-dependent ROC curves and (D) AUC. ROC=Receiver operating characteristic; AUC=Area under curve; SN=Substantia nigra; NM=Neuromelanin; FW=Free water; LC/LsC=Locus coeruleus/subcoeruleus.

When putamen DaT and SN NM were simultaneously considered as predictors, putamen DaT showed a slightly attenuated relationship with phenoconversion risk [HR (95%CI)=2.95 (1.4-6.3), χ^2^(1)=7.9, P=0.005] whereas the relationship with SN NM was rendered non-significant (P=0.16). There was no interaction effect between putamen DaT and SN NM (P=0.63).

### Free water in the substantia nigra relates to phenoconversion risk, but not when adjusting for neuromelanin

Higher SN FW was associated with higher phenoconversion risk [**Table 2**; **Figure 2A**; HR (95%CI)=1.54 (1.06-2.24), χ^2^(1)=5.1, P=0.025]. SN FW showed modest performance in ranking patients based on time to phenoconversion [**Figure 2B**; C-index=0.66] and in identifying converters [**Figure 2C-D**; iAUC=0.69].

When SN FW and SN NM were simultaneously considered as predictors, SN NM [HR (95%CI)=2.4 (1.2-4.7), χ^2^(1)=6.7, P=0.01] showed an association with phenoconversion risk whereas SN FW did not (P=0.11). There was no interaction effect between SN NM and SN FW (P=0.41). Similarly, when putamen DaT was included as a predictor, putamen DaT showed an association with phenoconversion risk (HR (95%CI)=3.2 (1.7-6.1), χ^2^(1)=12.3, P<0.001) whereas SN FW did not (P=0.96). There was no interaction effect between SN FW and putamen DaT (P=0.62).

### Neuromelanin in the locus (sub)coeruleus captures additional variability in phenoconversion risk

Lower LC/LsC NM was associated with higher phenoconversion risk [**Table 2**; **Figure 2A**; HR (95%CI)=2.1 (1.2-3.7), χ^2^(1)=6.5, P=0.011]. LC/LsC NM showed modest performance in ranking patients based on time to phenoconversion [**Figure 2B**; C-index=0.63] and in identifying converters [**Figure 2C-D**; iAUC=0.68].

When simultaneously included as predictors, both SN NM [HR (95%CI)=2.8 (1.4-5.6), χ^2^(1)=9.1, P=0.003] and LC/LsC NM [HR (95%CI)=2.3 (1.1-4.8), χ^2^(1)=4.3, P=0.037] retained their associations with phenoconversion risk. There was no interaction effect between SN NM and LC/LsC NM (P=0.98).

### Exploratory assessments of phenoconversion as a function of composite imaging scores

All three composite metrics were associated with phenoconversion risk. Lower Putamen DaT+SN NM was associated with higher phenoconversion risk [**Table 2**; **Figure 2A**; HR (95%CI)=3.4 (1.7-6.7), χ^2^(1)=12.5, P<0.001]. Putamen DaT+SN FW was associated with higher phenoconversion risk [HR (95%CI)=2.3 (1.4-3.7), χ^2^(1)=10.5, P=0.001]. Lower SN NM+LC/LsC NM was associated with higher phenoconversion risk [HR (95%CI)=3.0 (1.7-5.5), χ^2^(1)=13.5, P<0.001].

Putamen DaT+SN NM showed excellent performance in ranking patients based on time to phenoconversion [**Figure 2B**; C-index=0.81] and in identifying converters [iAUC=0.85]. Putamen DaT+SN FW showed good performance in ranking patients based on time to phenoconversion [C-index=0.77] and excellent performance in identifying converters [iAUC=0.80]. SN NM+LC/LsC NM showed good performance in ranking patients based on time to phenoconversion [C-index=0.77] and excellent performance in identifying converters [iAUC=0.81].

### Associations with Parkinson’s disease-specific phenoconversion

When assessing relationships with Parkinson’s disease-specific phenoconversion, both putamen DaT [HR (95%CI)=2.6 (1.8-5.7), χ^2^(1)=5.6, P=0.018] and SN NM [HR (95%CI)=2.6 (1.9-5.6), χ^2^(1)=5.7, P=0.017] were associated with risk. SN FW showed a trend towards an association with risk [HR (95%CI)=1.6 (0.38-2.60), χ^2^(1)=3.8, P=0.051], as did LC/LsC [HR (95%CI)=1.9 (0.94-1.06), χ^2^(1)=3.2, P=0.073].

### Imaging-based abnormality classifications capture phenoconversion risk

As a complementary approach to the reliance on continuous imaging predictors for the group-level analyses above, risk of phenoconversion in iRBD was additionally estimated based on binary classifications of abnormality (normal vs. abnormal). Imaging-based abnormality cut-offs were derived independently from patients with Parkinson’s disease and healthy controls. These cut-offs were subsequently applied to imaging data from each iRBD patient to classify them as normal (i.e. comparable to healthy control values) or abnormal (i.e., comparable to values from patients with Parkinson’s diseases). Putamen DaT showed excellent discriminative performance between patients with Parkinson’s disease and healthy controls (**Table 3**). When the same cut-off was applied to iRBD patients, 29% were classified as abnormal [**Table 3**; **Figure 3A**; AUC (95% CI)=0.98 (0.97-1), cut-off=2.85]. Putamen DaT abnormality was associated with the highest phenoconversion risk [**Figure 3B**; HR (95%CI)=8.4 (2.9-24.0), χ^2^(1)=15.6, P<0.001]. SN NM showed good discriminative performance, yielding a cut-off that classified 29% of iRBD patients as abnormal [**Figure 3C**; AUC (95% CI)=0.78 (0.70-786), cut-off=110.78]. SN NM abnormality was associated with higher phenoconversion risk [**Figure 3D**; HR (95%CI)=5.7 (2.0-16.4), χ^2^(1)=10.4, P=0.001]. SN FW showed modest discriminative performance, yielding a cut-off that classified 40% of iRBD patients as abnormal [**Figure 3E**; AUC (95%)=0.64 (0.56-0.72), cut-off=0.243]. SN FW abnormality was associated with higher phenoconversion risk [**Figure 3F**; HR (95%CI)=2.5 (1.0-6.0), χ^2^(1)=4.1, P=0.044]. LC/LsC NM showed the lowest discriminative performance, yielding a cut-off that classified 79% of iRBD patients as abnormal [**Figure 3E**; AUC (95%CI)=0.60 (0.51-0.69), cut-off=126.48]. LC/LsC NM abnormality showed no relationship with phenoconversion risk (**Figure 3H**; P=0.44).

**Table 3.** Imaging-based abnormality classifications and their relationship to iRBD phenoconversion risk.

| Predictor | AUC<sup>1</sup> | Cut-off<sup>1</sup> | N (% abnormal)<sup>2</sup> | HR (95% CI)<sup>2</sup> | $\chi^2(1)$<sup>2</sup> | P-value<sup>2</sup> |
| --- | --- | --- | --- | --- | --- | --- |
| Putamen DaT | 0.96 | 2.847 | 41 (29%) | 8.4 (2.9-24.0) | 15.6 | <0.001 |
| SN NM | 0.70 | 110.775 | 41 (29%) | 5.7 (1.98-16.4) | 10.4 | 0.001 |
| SN FW | 0.64 | 0.243 | 47 (40%) | 2.49 (1.0-6.0) | 4.1 | 0.044 |
| LC NM | 0.60 | 126.475 | 47 (79%) | 1.56 (0.5-4.7) | 0.6 | 0.44 |

<sup>1</sup>Discriminative performance of imaging metrics and associated cut-off values derived from a logistic regression analysis of Parkinson's disease patients versus healthy controls.
<sup>2</sup>Results obtained after applying the cut-off to distinguish abnormal and normal iRBD patients.

**Figure 3.**
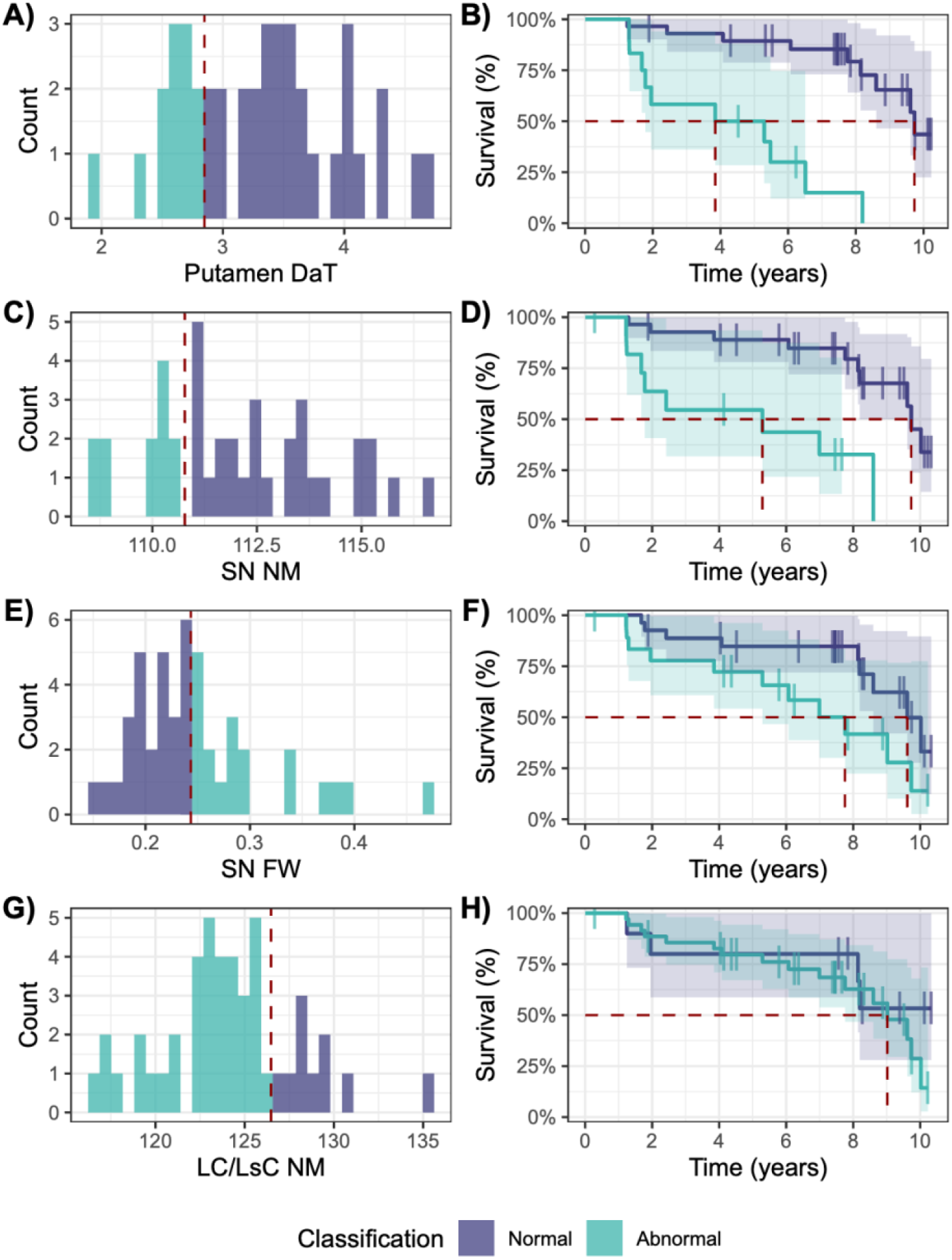
Imaging-based abnormality classifications in iRBD patients. (A) Abnormal putamen DaT counts and (B) association with phenoconversion risk. (C) Abnormal SN NM counts and (D) association with phenoconversion risk. (E) Abnormal SN FW counts and (F) association with phenoconversion risk. (G) Abnormal LC/LsC NM counts and (H) association with phenoconversion risk. Vertical dotted red lines in histograms denote cut-off values for abnormality classification (left) and median disease-free time in Kaplan-Meier plots separately for normal and abnormal groups. SN=Substantia nigra; NM=Neuromelanin; FW=Free water; LC/LsC=Locus coeruleus/subcoeruleus.

Correspondences between imaging-based abnormality classifications were relatively limited. Between putamen DaT and SN NM, cut-off-based abnormality classifications matched in 62% of cases. Between putamen DaT and SN FW, 67% of cases matched. Between SN NM and SN FW, 58% of cases matched.

## Discussion

We investigated the relationship between nigro-striatal deficits and risk of phenoconversion to overt synucleinopathies in a single-center cohort of polysomnography-confirmed iRBD patients who were monitored clinically over a period of 11 years. Out of 56 iRBD patients, 24 phenoconverted (14 Parkinson’s disease; 8 dementia with Lewy bodies; 2 multiple system atrophy). Our results replicate the well-established relationship between phenoconversion risk and putamen DaT.^2,7–11,17,71,72^ We extend this finding by showing that phenoconversion risk relates similarly to substantia nigra (SN) neuromelanin (NM) and free water (FW), thereby demonstrating involvement of the broader nigro-striatal dopaminergic system. In addition, we observed an association between LC/LsC NM and phenoconversion risk independently of SN NM, suggesting potentially distinct contributions from the dopaminergic and noradrenergic systems.

### Nigro-striatal deficits relate to phenoconversion risk in iRBD

We provide converging evidence that deficits in the nigro-striatal dopaminergic system contribute to phenoconversion risk in iRBD. Previous studies have demonstrated a clear relationship between elevated phenoconversion risk and reduced putamen DaT.^2,7–11,17,71,72^ However, it remains unclear to what extent the loss of nigral dopaminergic neurons contribute to this relationship, in addition to the decline in dopaminergic function at the level of nigrostriatal terminals. By showing that reduced SN NM and FW are associated with higher phenoconversion risk, we provide support for the view that pre-synaptic dopaminergic impairment and nigral cell loss contribute similarly to iRBD phenoconversion.

Our findings indicate that SN NM captures variability in phenoconversion risk, but that this variability is largely shared with putamen DaT, which emerged as a relatively stronger predictor. This aligns with research showing that striatal dopamine depletion typically exceeds the extent of SN cell loss, particularly during early disease stages.^22,23^ Consistently, when correcting for putamen DaT, the relationship between SN NM and phenoconversion risk was substantially weakened. Additionally, our composite metric combining putamen DaT and SN NM yielded relatively minor improvements in HRs, further emphasizing that SN NM and putamen DaT capture overlapping portions of variability in phenoconversion risk.

Our observation that SN NM and putamen DaT explain similar variability in phenoconversion risk opens new possibilities for monitoring phenoconversion risk in iRBD non-invasively, using MRI-based techniques, rather than nuclear imaging, which could help cut costs and reduce patient burden in clinical trials. To facilitate this development, we derived cut-offs from and independent cohort of patients with Parkinson’s disease and healthy controls to classify iRBD patients using abnormality cut-offs derived. Putamen DaT and SN NM cut-offs identified similar proportions of iRBD patients at risk of imminent phenoconversion. However, a caveat to this finding is that the overlap between the at-risk groups was only partial, suggesting that the two metrics may identify partly distinct sub-populations of patients. While a lack of classification overlap may relate to underlying biological differences, such as the trajectory of pathological propagation,^29,73^ it may also relate differences in precision of distinct imaging metrics. Additional verification in independent sample is therefore required.

### The relationship between phenoconversion risk and dopaminergic cell loss in the substantia nigra

We provide converging evidence for a relationship between phenoconversion in iRBD and dopaminergic cell loss in the SN using two well-established imaging metrics in research on synucleinopathies: NM and FW.^15^ A relationship between iRBD phenoconversion and SN cell loss has previously been demonstrated using the FW metric.^18^ However, since FW is sensitive primarily to increases in extracellular water,^50^ it remains unclear how phenoconversion in iRBD relates specifically to loss of dopaminergic cells, as opposed to more general neurodegeneration in the midbrain. In contrast, NM-MRI contrast has been associated with the presence of NM-iron complexes in the SN^74–76^ and is thought to reflect degeneration of dopaminergic cells more directly.^77^ Our observation that SN NM relates to phenoconversion risk in iRBD, even when adjusting for more general neurodegenerative processes captured by FW, therefore provides compelling evidence for the involvement of dopaminergic cell loss in iRBD phenoconversion.

The increased sensitivity of NM-MRI with respect to dopaminergic cell loss may explain why SN NM emerged as a more sensitive predictor of phenoconversion compared to SN FW. However, additional factors may have contributed, such as the spatial specificity of the regions of interest that were used to extract metrics from the SN. Our NM pipeline involved automated segmentations of the whole SN in native space.^45^ In contrast, our FW pipeline relied on a box-shaped region of interest manually positioned in the posterior segment of the SN in MNI space.^48,49^ Consequently, noise due to interindividual differences in region of interest placement and degree of partial volume effects following may be higher in our FW metric, which could contribute to its relatively lower sensitivity with respect to phenoconversion risk.

### Dopaminergic and noradrenergic systems relate independently to phenoconversion risk

Synucleinopathies display prominent deficits in noradrenergic neurotransmission.^28,35^ These deficits have also been observed in iRBD,^30–32,78^ suggesting that they may herald the transition from pre-symptomatic to clinically overt neurodegenerative disease. We provide evidence for this by demonstrating that phenoconversion risk in iRBD is increased when LC/LsC NM signal is reduced. There are multiple routes along which LC/LsC degeneration may contribute to phenoconversion.^35^ Degeneration of the LC/LsC and loss of noradrenaline sensitizes dopaminergic neurons in the SN to damage.^26^ Moreover, LC/LsC degeneration impairs the capacity to integrate information across distinct functional brain networks,^79,80^ which could disrupt compensatory mechanisms that otherwise stabilize behavior against pathological decline in the nigro-striatal system.^51,81,82^

Alternatively, LC/LsC NM may relate to phenoconversion simply because its trajectory closely resembles that of the SN.^28^ Our finding that LC/LsC and SN degeneration relate independently to phenoconversion argues against this view, and may be taken as preliminary evidence for distinct contributions of noradrenergic and dopaminergic deficits. This fits with recent work showing differential contributions of LC/LsC and SN degeneration to cognitive deficits in Parkinson’s disease.^83^ However, it should be noted that NM-MRI cannot distinguish the noradrenergic and non-noradrenergic nuclei comprising the LC/LsC. Thus, a mechanistic role for noradrenaline cannot be concluded solely from the results of this study. Regardless, our findings demonstrate that the complementary information of SN and LC/LsC NM may be leveraged to improve tracking of phenoconversion in iRBD. This is supported by our exploratory analysis of composite scores, showing that the combination of SN and LC/LsC NM yielded similar HRs as putamen DaT in isolation, providing further motivation for considering NM-MRI as a potential alternative to ^123^I-FP-CIT SPECT when investigating phenoconversion risk in iRBD.

### Strengths, limitations and interpretational issues

In this unique single-centre study, we employed a combination of multimodal imaging, comprising ^123^I-FP-CIT SPECT, NM-MRI, and diffusion-weighted MRI acquired from the same iRBD patients, together with a sufficiently long follow-up duration to observe rising phenoconversion rates. Our observed phenoconversion rates and diagnostic distributions largely converged with previous cohort studies and meta-analyses,^2,3,84,85^ as did our findings on the relationship between phenoconversion and striatal dopamine.

Our study has some limitations. Findings of similarities in prognostic associations between NM-MRI and ^123^I-FP-CIT SPECT should not be interpreted as demonstrating equivalent readiness for multi-centre trial stratification. Cross-site assessments of reproducibility and transportability of derived cut-offs are still needed to account for sequence-and scanner-dependency of NM-MRI measurements.^74,86^ In contrast, ^123^I-FP-CIT SPECT has well-established international procedural standards,^87^ clinically deployed semiquantification software,^88^ and multi-centre normative databases,^89^ which are not yet available for NM-MRI.

The relatively small sample size prevented us from investigating disease-specific effects. Further research in larger, multi-center samples will be required to determine whether SN and LC/LsC degeneration, as measured by NM-MRI, differentially tracks phenoconversion to Parkinson’s disease or dementia with Lewy bodies.

Imaging-based abnormality classifications were based on reference values from a sample of patients with Parkinson’s disease. Given that iRBD also predisposes patients to dementia with Lewy bodies and multiple system atrophy, results from these analyses should be interpreted with caution. In addition, as has been observed in previous studies,^2,3,84,85^ our sample showed a relatively greater prevalence of converters to Parkinson’s disease, rather than dementia with Lewy bodies, which imposes limits on the generalizability of our findings.

## Conclusion

We provide converging evidence that dopaminergic deficits in the nigro-striatal system herald phenoconversion in iRBD and highlight potentially distinct contributions from the noradrenergic locus coeruleus/subcoeruleus. These findings contribute to improved monitoring of phenoconversion risk in iRBD and present opportunities to use neuromelanin-sensitive MRI for stratification and the definition of intervention targets in neuroprotective trials.

## Data availability

Data and code supporting the findings of this study are available upon reasonable request.

## Acknowledgements

The authors would like to thank Abel Grine, for assisting in the reconstruction of ^123^I-FP-CIT SPECT images, and the ICEBERG Study Group.

## Supplementary material

Supplementary material is available at *Brain* online.

## Supplementary materials

### Effects of data missingness in Cox proportional hazards regression analyses

#### Complete-case analysis

Cox proportional hazards regression was performed in a subset of participants with complete-case data. Sample size was restricted to participants without missing data for putamen DaT, SN NM, SN FW, and LC/LsC NM. This led to a total sample size of 29 participants, out of whom 15 phenoconverted (9 PD; 5 DLB; 1 MSA).

Lower putamen DaT was associated with higher phenoconversion risk [HR (95%CI)=2.9 (1.6-5.4), χ^2^(1)=14.7, P<0.001]. Putamen DaT showed excellent performance in ranking patients based on time to phenoconversion [C-index=0.81] and in identifying converters [iAUC=0.79].

Lower SN NM was similarly associated with higher phenoconversion risk [HR (95%CI)=2.1 [1.1-4.4], χ^2^(1)=4.5, P=0.034]. SN NM showed modest performance in ranking patients based on time to phenoconversion [C-index=0.68] and in discriminating converters from non-converters [iAUC=0.66].

SN FW and LC/LsC NM showed no association with phenoconversion risk (P=0.19 and P=0.18, respectively).

#### Multiple imputation analysis

Multiple imputation with predictive mean matching^1,2^ was used to perform Cox proportional hazards regression across the full sample of 56 participants. The number of imputed data sets were computed separately for each outcome of interest, with a minimum boundary set to 20.^3^ Estimates were pooled according to Rubin’s rules.^4^ For these analyses, discriminative performance was not considered.

Lower putamen DaT was associated with higher phenoconversion risk [HR (95%CI)=3.0 (1.7-5.5), t(17.7)=3.9, P<0.001, number of imputations=30]. Lower SN NM was similarly associated with higher phenoconversion risk [HR (95%CI)=2.6 [1.3-5.1], t(16.6)=3.0, P=0.008, number of imputations=78]. Higher SN FW was associated with higher phenoconversion risk [HR (95%CI)=1.5 (1.0-2.3), t(19.6)=2.2, P=0.04, number of imputations=20]. Lower LC/LsC NM was associated with higher phenoconversion risk [HR (95%CI)=2.0 (1.1-3.6), t(17.7)=2.5, P=0.023, number of imputations=30].

